# Rapid colorimetric detection of COVID-19 coronavirus using a reverse transcriptional loop-mediated isothermal amplification (RT-LAMP) diagnostic platform: iLACO

**DOI:** 10.1101/2020.02.20.20025874

**Authors:** Lin Yu, Shanshan Wu, Xiaowen Hao, Xuelong Li, Xiyang Liu, Shenglong Ye, Heng Han, Xue Dong, Xin Li, Jiyao Li, Na Liu, Jianmin Liu, Wanzhong Zhang, Vicent Pelechano, Wei-Hua Chen, Xiushan Yin

**Affiliations:** Applied Biology Laboratory, Shenyang University of Chemical Technology, 110142, Shenyang, China; Key Laboratory of Molecular Biophysics of the Ministry of Education, Hubei Key Laboratory of Bioinformatics and Molecular-imaging, Center for Artificial Intelligence Biology, Department of Bioinformatics and Systems Biology, College of Life Science and Technology, Huazhong University of Science and Technology, 430074 Wuhan, Hubei, China; SciLifeLab, Department of Microbiology, Tumor and Cell Biology. Karolinska Institutet, Solna 171 65, Sweden; College of Life Science, HeNan Normal University, 453007 Xinxiang, Henan, China; Pluri Biotech Co.Ltd, Xuzhou, 221001, China; Shenyang Center for Disease Control And Prevention, 110031, Shenyang, Liaoning, China; Biotech & Biomedicine Science (Shenyang)Co. Ltd, Shenyang, 110000, China; Nanog Biotech Co.Ltd, Shanghai, 200000,China; Biotech & Biomedicine Science (Jiangxi) Co. Ltd, Ganzhou, 341000, China

**Keywords:** isothermal amplification, COVID-19, LAMP, iLACO

## Abstract

The recent outbreak of a novel coronavirus SARS-CoV-2 (also known as 2019-nCoV) threatens global health, given serious cause for concern. SARS-CoV-2 is a human-to-human pathogen that caused fever, severe respiratory disease and pneumonia (known as COVID-19). By press time, more than 70,000 infected people had been confirmed worldwide. SARS-CoV-2 is very similar to the severe acute respiratory syndrome (SARS) coronavirus broke out 17 years ago. However, it has increased transmissibility as compared with the SARS-CoV, e.g. very often infected individuals without any symptoms could still transfer the virus to others. It is thus urgent to develop a rapid, accurate and onsite diagnosis methods in order to effectively identify these early infects, treat them on time and control the disease spreading. Here we developed an isothermal LAMP based method-iLACO (*i*sothermal <u>LA</u>MP based method for <u>CO</u>VID-19) to amplify a fragment of the ORF1ab gene using 6 primers. We assured the species-specificity of iLACO by comparing the sequences of 11 related viruses by BLAST (including 7 similar coronaviruses, 2 influenza viruses and 2 normal coronaviruses). The sensitivity is comparable to Taqman based qPCR detection method, detecting synthesized RNA equivalent to 10 copies of 2019-nCoV virus. Reaction time varied from 15-40 minutes, depending on the loading of virus in the collected samples. The accuracy, simplicity and versatility of the new developed method suggests that iLACO assays can be conveniently applied with for 2019-nCoV threat control, even in those cases where specialized molecular biology equipment is not available.

## Introduction

In December 2019, the first case of cluster unidentified pneumonia was found in Wuhan, named 2019-Novel Coronavirus (2019-nCoV), a beta subtype, highly homologous with SARS(1). As of February 18, 2020, there were in total 75,201 confirmed cases worldwide. With the rapid spreading speed of R0 3.28, there is a heavy pressure for hospital assistant diagnosis, as some infected individuals without any symptoms could still transfer the virus to others(2). Thus, there is an urgent need to develop a diagnostic method that is sensitive, accurate, rapid and low-cost to screen infected individuals, facilitate proper isolation and help controlling the disease spread.

For RNA virus infections, especially acute respiratory infection, probe coupled RT-qPCR from respiratory secretions is routinely used to detect causative viruses (3–9). This method has been widely used by Center for Disease Control and Prevention and other relevant departments worldwide. Recently RT-qPCR based methods were developed and applied worldwide by multiple research and disease control centers (10). However, RT-qPCR has many limitations such as the need for high purity samples and the access to expensive laboratory instruments, as well as requiring long reaction times (at least 2 h). In addition, RT-qPCR needs trained personnel and sophisticated facilities for sample processing. These disadvantages limit its practical application in many cases, and thus can delay the required rapid prescription and administration of antiviral agents to patients.

Loop -mediated isothermal amplification (LAMP) is a technology that provides nucleic acid amplification in a short time using 4 to 6 specially designed primers and a DNA polymerase with chain displacement activity (Bst) under a constant temperature (60-65□)(11, 12). Particularly relevant for this application, is that LAMP can be combined with reverse transcription (*i*.*e*. RT-LAMP), were both reverse transcription and amplification occur simultaneously allowing the direct detection of RNA (13–17). This system, can be coupled with a pH indicator present in the reaction mix allowing readout of the amplification reaction by change in color (18). Therefore, LAMP has the potential to offer an easy diagnostic test where the result could be directly observe the color change with the naked eye.

The present study describes an isothermal LAMP based method-iLACO (*i*sothermal <u>LA</u>MP based method for <u>CO</u>VID-19), for rapid colorimetric detection of 2019-Novel Coronavirus.

## Materials and Methods

### Clinical samples

Respiratory samples were obtained from patients sent to the Shenyang CDC and tested by the RT-qPCR kit. RNA was further diluted for iLACO development and evaluation.

### RNA extraction

RNA was extracted from clinic samples with COVID-19 virus infection during the epidemic in Shenyang in 2020 with the Thermo nuclear acid extraction workstation in the P3 laboratory of CDC Shenyang. Before the experiment, the concentration of virus RNA sample was measured with Qubit 3.0 (Thermo Fisher).

### Reverse Transcription

COVID-19 virus RNA was reverse transcribed using random primer with SuperScript® III Reverse Transcriptase First-Strand Synthesis System (Invitrogen, USA). The reverse transcription reaction was carried out at 50°C for 1 h. The cDNA obtained was used as the detection sample of RT-LAMP reaction as well.

### Real-time reverse-transcription PCR

Two kits were used in parallel to validate the positive samples (BGI and Biotech & Biomedicine Science (Shenyang)). Briefly, a 20 µl reaction volume containing 5μl RNA template, 13 μl master mix, 2 μl primer and probe mix. The reverse transcription reaction was performed at 50° for 5 minutes, the initial denaturation was 94° for 2 minutes, 40 cycles of denaturation at 94° for 5s and annealing at 55° for 10s. The PCR apparatus used in this research were CFX96 TouchTM Real-Time PCR Detection System (Bio-Rad, Hercules, CA, USA) and ABI 7500 Real-time PCR system.

### iLACO development

To develop the iLACO, we first use the commercialized WarmStart master mix kit from NEB combined with 10 panel of designed primers. A 20 µl reaction contained 10µl WarmStart Colorimetric LAMP 2X Master Mix, 2µl primer mix, 1µl RNA/cDNA sample as positive reaction and 7µl DEPC-treated water. For the negative control group, we replaced the template RNA/cDNA with DEPC-treated water. Mixed reactions were all performed in PCR thermo-cycler or water bath incubation at 65 °C for 20-40 minutes. Positive reactions resulted in a color change of phenol red pH indicator from pink to yellow due to decreased pH in the presence of extensive DNA polymerase activity. However, the negative reaction still kept pink. Later we prepared all the solutions with individual components to optimize the reaction solution, also with different dyes for color judgement sensitivity test.

### Ethical statement

Sample collection and analysis of samples were approved by the local CDC. The internal use of samples was agreed under the medial and ethical rules of each participating individuals.

### Primer design and analysis

RT-LAMP primers were designed by the online software Primer Explorer V5 (http://primerexplorer.jp/lampv5e/index.html). For the primer sets, we used the online tool Primer-BLAST (https://www.ncbi.nlm.nih.gov/tools/primer-blast/index.cgi?LINK_LOC=BlastHome) (19) to detect whether the primer sequence is specific to the human genome and retain the primers that all 3 pairs primers are specific. Then compare the primer region sequence with other viruses’ genome by the online tool BLAST (https://blast.ncbi.nlm.nih.gov/Blast.cgi?PROGRAM=blastn&PAGE_TYPE=BlastSearch&LINK_LOC=blasthome)(20) to calculate the homology.

The final set of primers used in this research for iLACO are shown in Table 1. The concentration of each primer in primer mix were as followed: 0.2 µM of each outer primer (F3 and B3), 1.6 µM of each inner primer (FIP and BIP), 0.4 µM of each loop primer(LF and LB)(21).

**Table 1.**
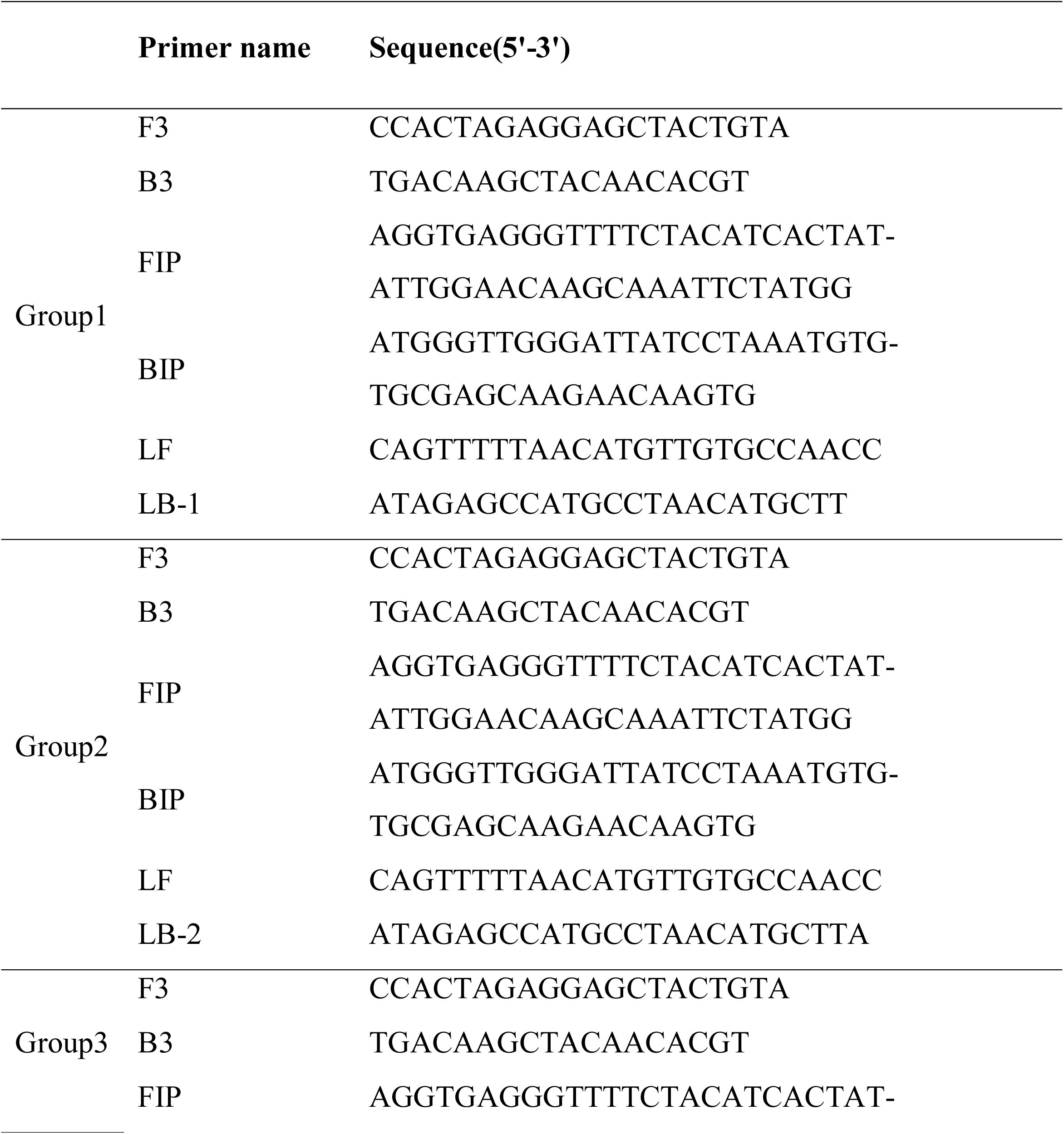
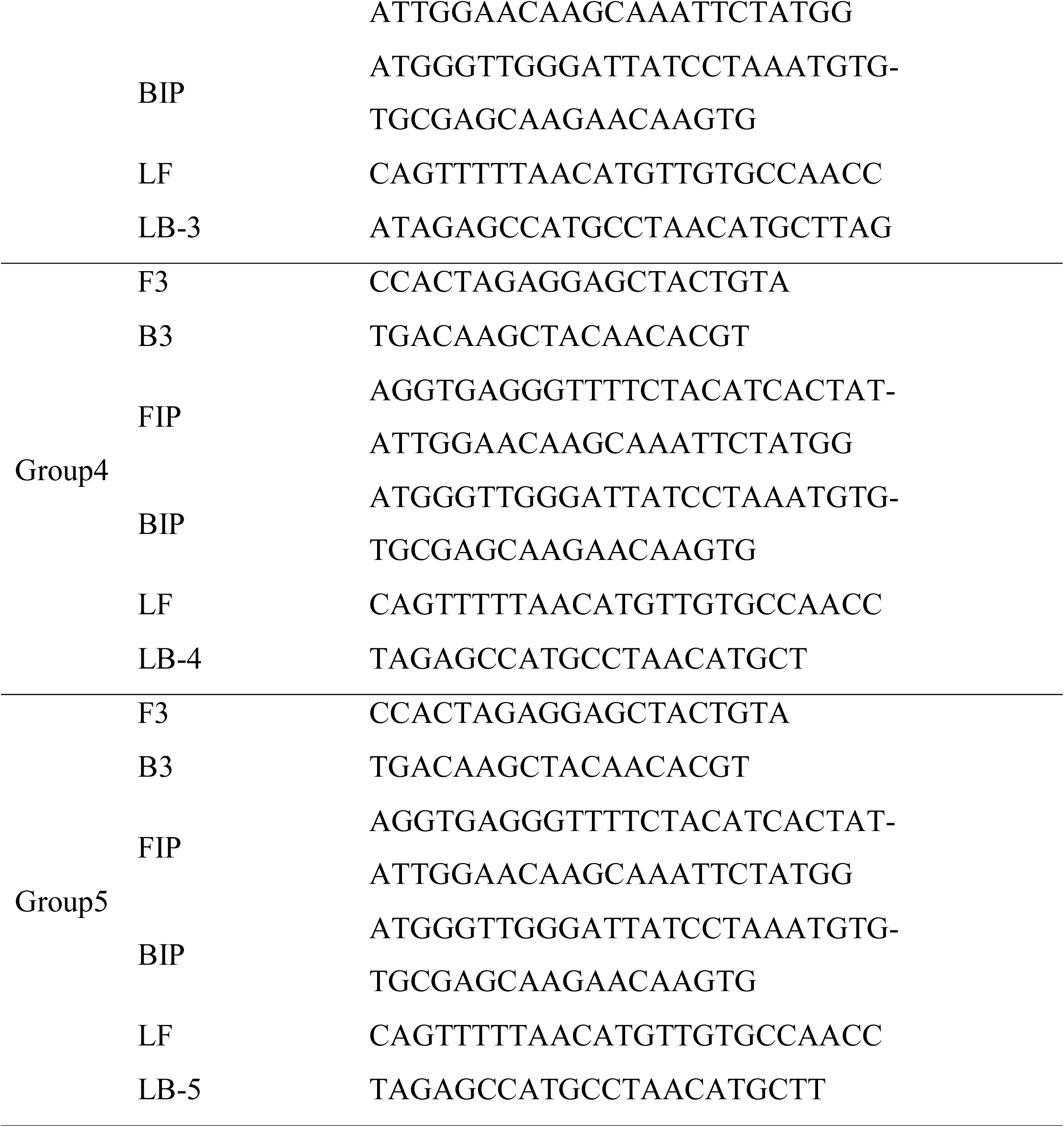
Primers used for RT-LAMP for COVID-19 detection.

## Results

### Primer region mapping

We selected a region of the ORF1ab as target region that is currently used for RT-qPCR detection approaches (Fig. 1A), and used the online software Primer Explorer V5 (http://primerexplorer.jp/lampv5e/index.html) to design the RT-LAMP primers. After the specificity analysis, we retained one primer set with several pairs of loop primers (Table 1, see Materilas and Methods for details); their genomic locations on the virus genome can be found in Fig.1B (see also Table 1). To assure the primer specificity, we compared their sequences with other viruses genome (include 7 similar coronaviruses, 2 influenza viruses, and 2 other coronaviruses)(22) by BLAST and found that the sequences were not similar to most viruses’ sequences we have chosen (Fig. 1C).

**Figure. 1.**
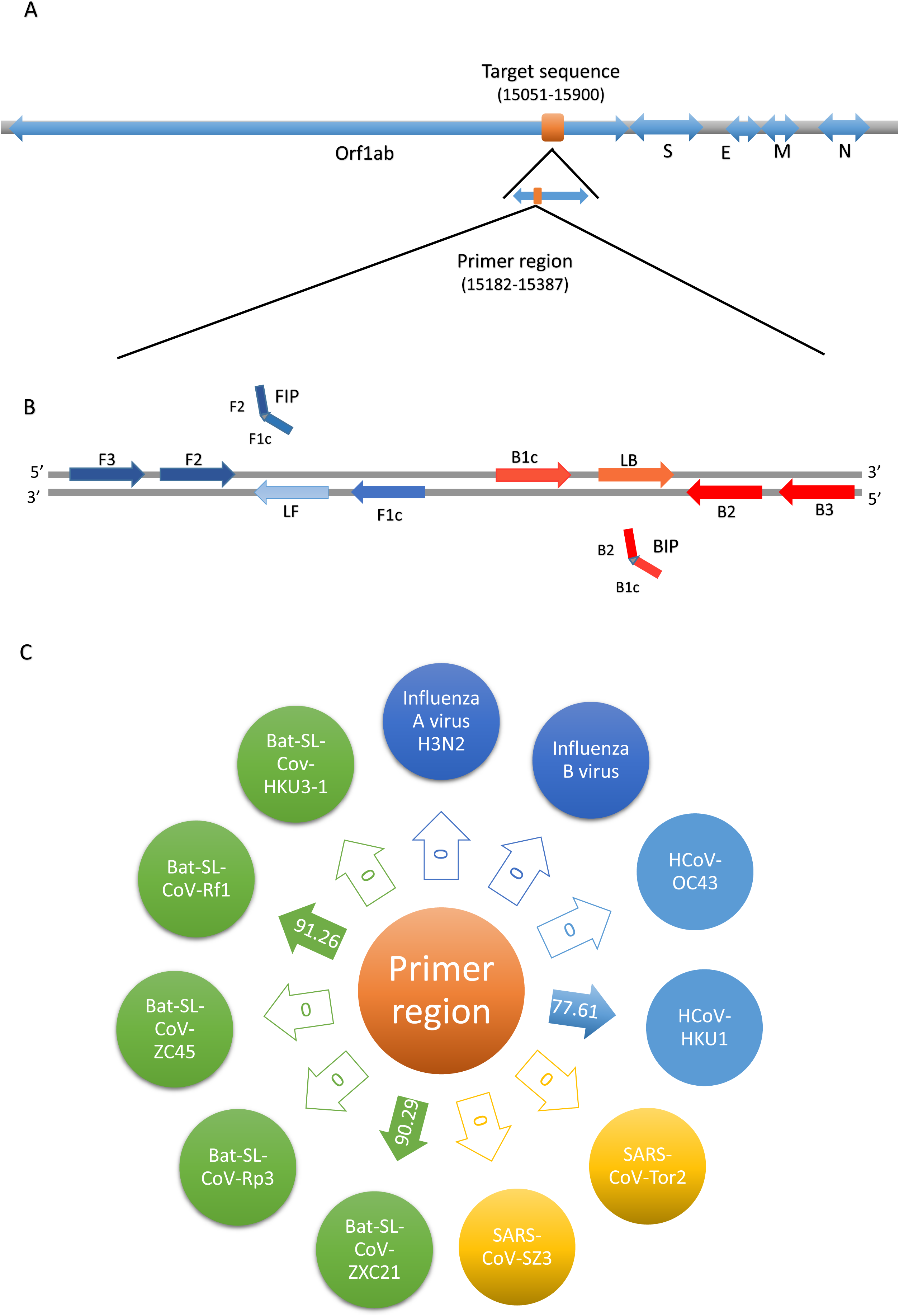
Primer mapping and homology of primer region sequence. A: The position of the target sequence on the complete SARS-CoV-2 genome sequence. B: The sites of primers. The 3 pairs primers include two inner primers (FIP/BIP), two outer primers (F3/B3) and two loop primers (LF/LB). C: Homology analysis with other viruses. The distance (percent identity) was calculated by comparing the primer region sequence with other viruses’ genome by BLAST software.

### Establishment and optimization of iLACO

We used one SARS-CoV-2 positive patient respiratory RNA sample validated by RT-qPCR to set up the iLACO assay. We confirmed that after 20 minutes of incubation in a thermoblock, using the designed RT-LAMP primers the targeted gene was successfully amplified and a color change was observed in the reaction tubes. Specifically, a change in color from pink to light yellow indicated a positive reaction while negative reactions retained a pink color.

To optimize of RT-LAMP reaction, we used five group of primers with variable LB oligos 1-5 (Table 1). We evaluated their ability to produce an effective amplification of both RNA and cDNA samples with 15 minutes or 20 minutes of incubation time(Fig. 2A and Fig. 2B). After the incubation, in addition to the colorimetric change, a ladder of DNA with increasing size was observed with on an agarose gel confirming the expected DNA amplification (Fig. 2C). The 5 sets of primers were further tested for efficiency and the primer group 4 (containing LB4) was chosen for further optimization, which showed the best sensitivity (data not shown). We also optimized the reaction temperature to 65° degree, as we did not observe any positive signal using the recommended 72° degree incubation previously used for ZIKA virus detection by LAMP assay(21). iLACO showed similar performance when we compared the samples from SARS-CoV-2 RNA or cDNA, indicating the one-step isothermal amplification is sufficient and separated step for reverse transcription is not necessary.

**Figure. 2.**
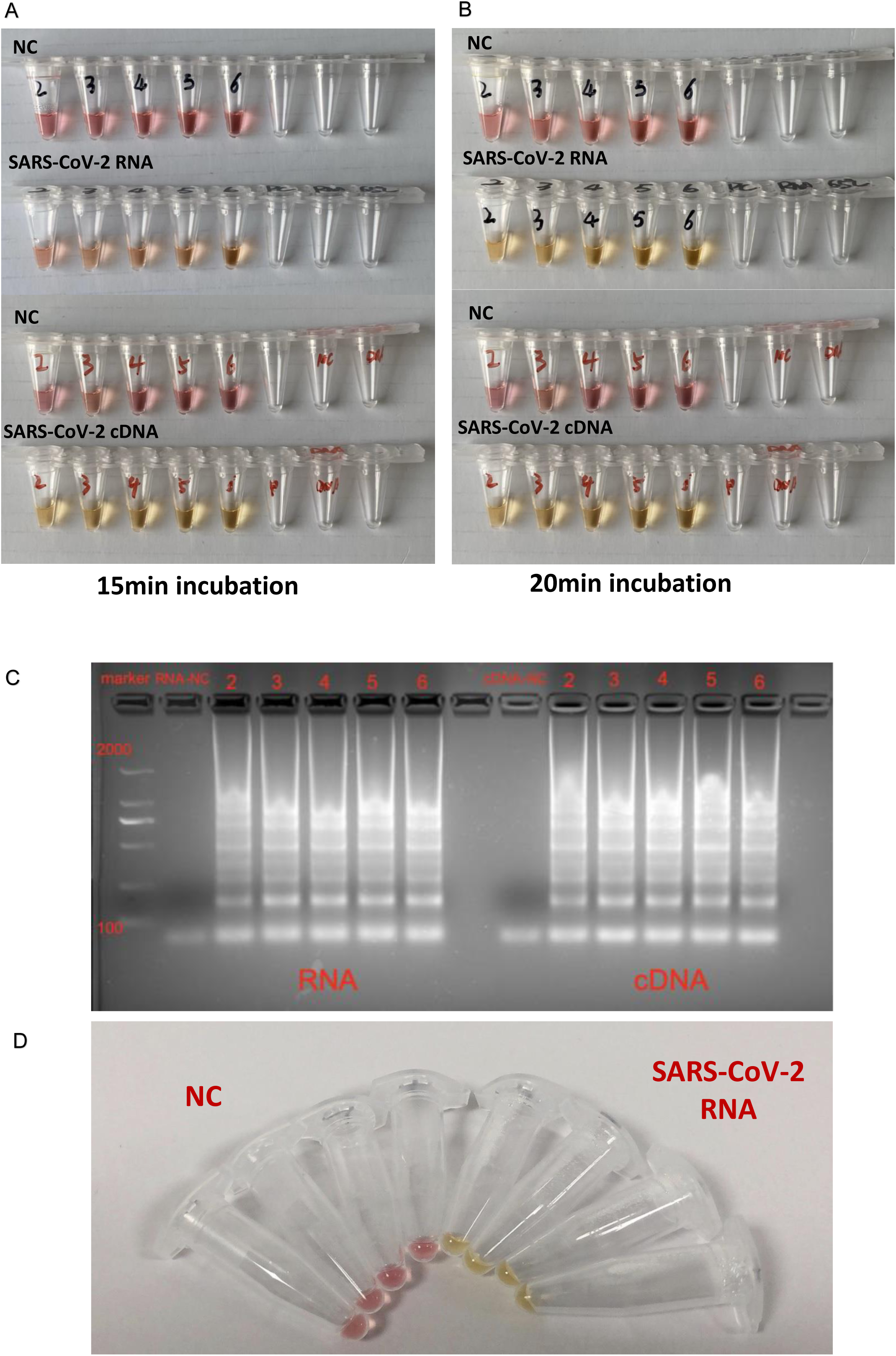
Time optimization of iLACO reaction of RNA and cDNA samples respectively. A and B: Five groups of specific primer sets to detect RNA and cDNA samples for 15 minutes (A) and 20 minutes (B). NC refers to the negative controls and the numbers in the tubes to the specific LB oligo (Table1). C: Amplification products checked by agarose gel electrophoresis. D: The signal of iLACO reaction in 1.5 ml tubes incubated with water bath or incubator. PC refers positive controls containing SARS-CoV-2 RNA.

In order to optimize the potential usage for field-based or bed-based detections, we checked the efficiency of iLACO in 1.5 ml tubes incubated with water bath or incubator. Twenty minutes of reaction is sufficient to detect the signal (Fig. 2D). We notice the signal is stronger compared with incubation using a thermocycler commonly used for PCR. The longer incubation time caused the water drops on the top lid of the tube. To avoid this, we added few drops of mineral oil after adding all the required solutions. This set up can be further optimised by using micro Pasteur Pipettes coupled with thermo cups for processing.

### Assay sensitivity compared with RT-qPCR

To check the detection limit of iLACO, we prepared multiple reactions containing serial dilutions of synthesized ORF1ab gene (from 1,000,000 to 10 copies). As shown in Figure 3, iLACO is very sensitive, and as low as 10 copies of ORF1ab gene were detected successfully (1:100000 dilution in Fig. 3A). Further, we compared iLACO with RT-qPCR assay with the diluted RNA sample from patient number 200202(10 times dilution, SARS-CoV-2 virus concentration around 17 copies/µl).. We observed the iLACO reaction time correlating well with RT-qPCR cycles. ie. 37 minutes color turning point correlated with 37 cycles (Fig. 3B and C). However, the 20 µl volume reaction time in 1.5 ml tubes incubated on wather bath are much fastercompared to RT-qPCR or samples processedin 0.2 ml tubes in PCR thermocycler, partially due to the increased heating volume and even surrounding temperature.

**Figure. 3.**
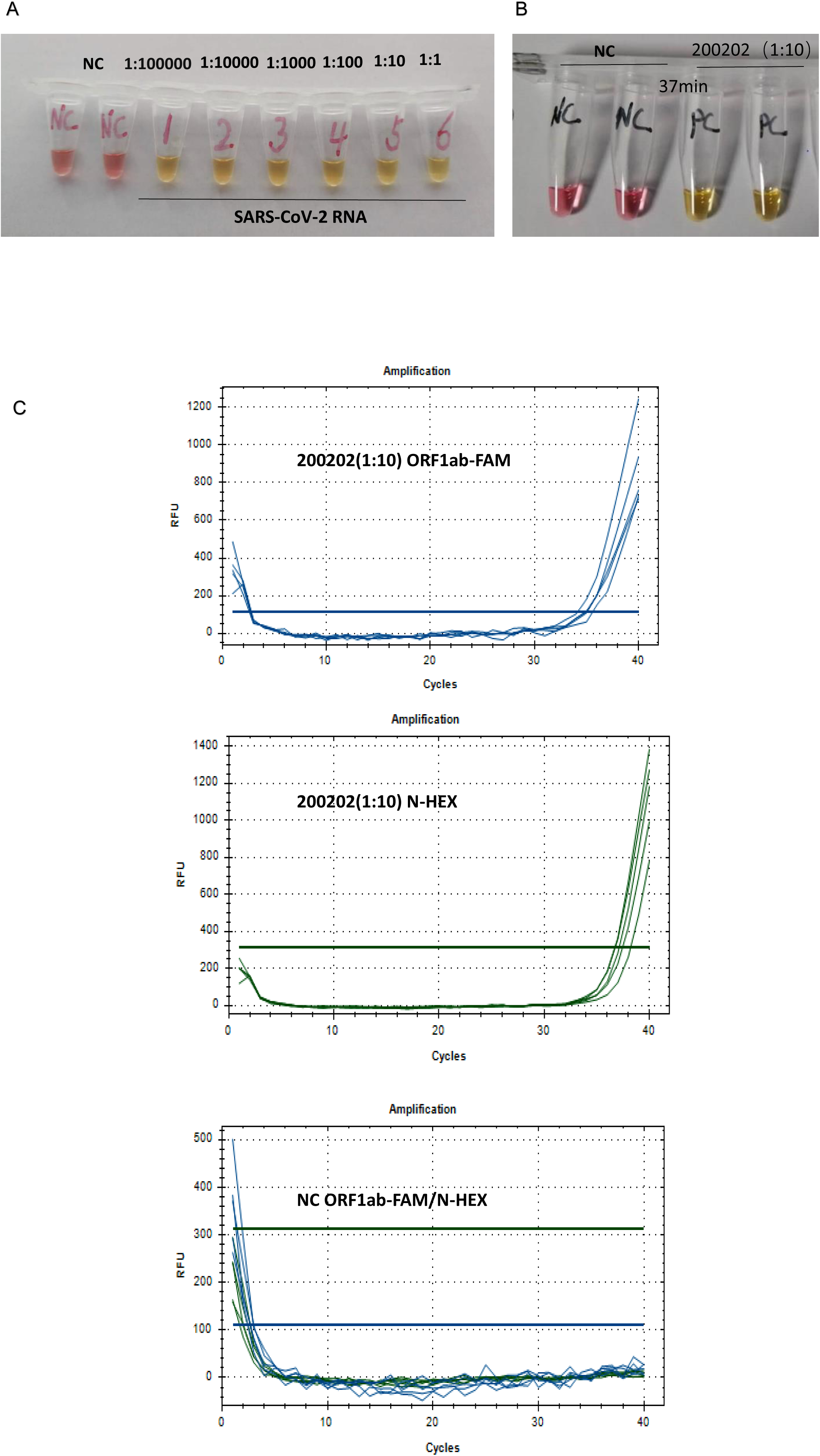
Sensitivity for iLACO compared with RT-qPCR assay. A: Series dilution of RNA for limitation evaluation. Note the 1:100000 equals 10 copies of synthesized ORF1ab RNA. B: SARS-CoV-2 RNA Sample 200202(1:10 diluted) A detected positive signal in 37 min. C: SARS-CoV-2 RNA Sample 200202(1:10 diluted) showed positive Ct value at 37. Two Taqman probes were used to target the ORF1ab and N gene, respectively.

### Detection of amplified product by alternative methods

To further expand the iLACO detection capability, we also checked the UV or Blue light stimulated fluorescence signal. We added SYBR green dye into the reaction mix prior to adding the sample. After 20 minutes incubation in 65 degree, the signal was observed with Gel imaging system (Fig. 4A). SYBR green in positive reactions was observed clearly in positive reaction. We also chose a new type of nucleic acid dye GeneFinder™, which has enhanced fluorescent signal and sensitivity. By exposing under Blue light, green fluorescence was observed clearly with naked eye in the positive reaction, whereas it remained original pink in the negative control (Fig. 4B).

**Figure. 4.**
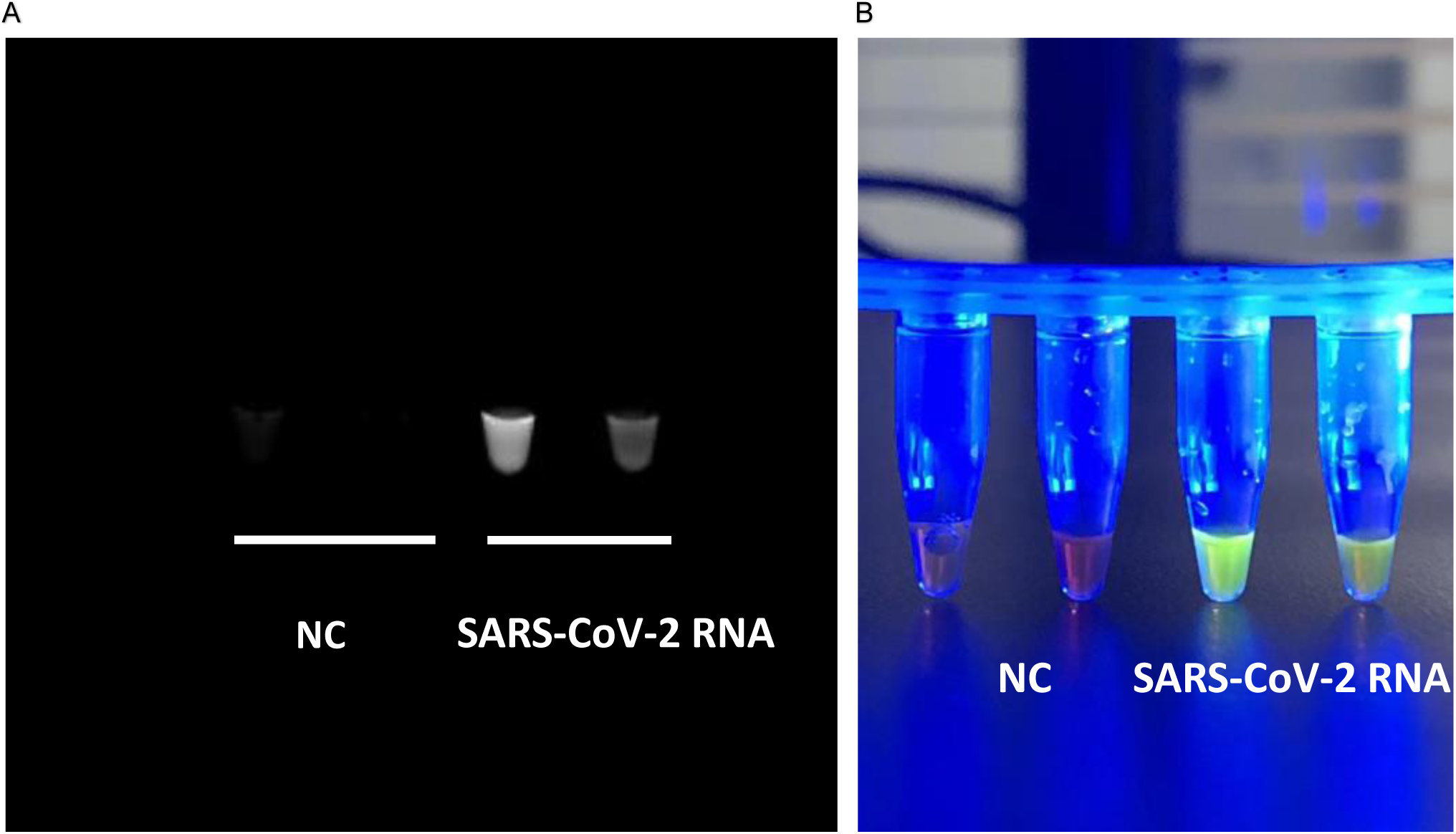
Fluorescent signal detected by UV and blue light for iLACO. A: The signal was detected with the gel imaging system after the SYBR green dye was added. NC and PC refers to the negative and COVID-19 positive control respectively. B: Positive signal was visible with the naked eye under blue light.

### Evaluation of the iLACO assay using clinical samples

Finally, we evaluated the iLACO with total 43 samples initially diagnosed with RT-qPCR during the epidemic in Shenyang in 2020. Results showed that 97.6% (42/43) of samples validated by qPT-PCR showed consistent signal after 40 min incubation with 2 µl sample loading(RNA concentration range 0.2-47 ng/µl). One sample kept unchanged color after 50 minutes, indicating the low dose of virus would cause false negative or spontaneous negative signal. To confirm this, we repeated this sample multiple times with random positive signal, confirming that this samples was close to the detection threshold using 2µl. Currently most the RT-qPCR reactions in China for SARS-CoV-2 test use 5 µl sample input. We thereby checked whether increasing the used sample volume would facilitate the detection. However, increasing the volume of the loaded sample to 5 µl RNA sample lead to variable results. This is most likely due to the presence of Tris or EDTA in the RNA dilution buffer when automatic RNA extraction workstation is used. This could be optimized by adjusting the concentration of used buffers. We recommend to use always a positive and a negative control sample resuspended in the same buffers used for patient RNA isolation.

## Discussion

The most popular methods for RNA virus detection so far are based on RT-PCR and/or RT-qPCR; while these methods only take a run time of 2-3 hours, they require special experimental apparatus, controlled working environment and well-trained personnel, which are often lacking in massive virus outbreaks such as the SARS or COVID-19. In this research, we developed an optimized iLACO that can rapid and sensitively detect COVID-19 virus RNA or cDNA samples based on LAMP isothermal amplification. The reaction time takes 20-30 minutes at 65°C. The result showed that by optimizing the concentration of primers, the total reaction time would be shorter for high dose virus carriers. iLACO detection method could not only detect the RNA of SARS-CoV-2 virus, but also worked on the cDNA samples.

The use of iLACO will facilitate the widely application of virus detection test, especially in developing countries with limited facilities, and when compared with other assays that are laborious, costly, and time-consuming(23).

Currently, there is a lack of specific drugs for the treatment of the novel coronavirus (COVID-19) pneumonia, so the early detection and following treatment is essential to control the spread of the disease. Therefore the detection of COVID-19 is of vital importance. The first version of iLACO with six primers could specifically identify eight distinct regions of the ORF1ab target. Our blast analysis suggest that the highly specific primer set design have a low chance for unspecific amplifications (false positives).

## Data Availability

COVID-19 samples were obtained from patients sent to the Shenyang Center for Disease Control And Prevention, and the nucleotide sequences were downloaded from NCBI (https://www.ncbi.nlm.nih.gov/) database.

## Acknowledgements

This work was funded by 2020 LiaoNing Provence Key Research Project (1580441949000), Ganzhou COVID-19 Emergency Research Project. VP is funded by the Swedish Research Council (VR 2016-01842), a Wallenberg Academy Fellowship (KAW 2016.0123), the Swedish Foundations’ Starting Grant (Ragnar Söderberg Foundation), Karolinska Institutet (SciLifeLab Fellowship, SFO and KI funds) and a Joint China-Sweden mobility grant from STINT (CH2018-7750). All the authors plan to make the reagents widely available to the community, primers used in this paper can be requested for free delivery through.

